# Skin in The Game: The Cost Consequences of Skin Cancer Diagnosis, Treatment and Care in Northern Ireland

**DOI:** 10.1101/2023.05.09.23289720

**Authors:** Ethna McFerran, Sarah Donaldson, Mark Lawler

## Abstract

**Background:** Skin cancer is a prevalent type of cancer in the UK. Its rising incidence and mortality rates are expected to result in substantial financial implications, particularly on diagnostic and treatment services for skin cancer management in Northern Ireland. Such anticipated disease increases underscore the need for prevention and control measures that should guide policymaking and planning efforts.

**Methods:** We conducted a retrospective cost study to measure the burden of skin cancer in Northern Ireland from a healthcare system perspective. Our data-driven model utilized bottom-up methodology ^1,2^ and reported 2018 costs using NHS reference unit costs (UK£) for skin cancer diagnosis and treatment patient pathways. Sensitivity analyses were performed, including varying diagnostic volumes by applying multipliers for benign cases and assuming a diagnostic conversion rate of 6.8%. An alternative chemotherapy regimen compliance rate was also examined at 75% as compared to base case. Proportional increases were projected based on future estimated increases of 9% and 28% in melanoma cases for diagnostic, treatment, and follow-up volumes specifically related to malignant melanoma.

**Results:** As of 2018, NICR recorded 4142 non-melanoma skin cancers (NMSC) and 423 malignant melanoma (MM) cases, averaging 17.5 new patients per trust weekly. The total costs for managing NMSC was £1,815,936, whereas that for MM skin cancer costs was £12,364,220, out of which £8,792,208 accounted for procurement, administration, and chemotherapy drug use. Healthcare providers spent a total of £17,024,115 on skin cancer care. Sensitivity analysis suggest diagnostic cost would either reduce by £781k to £3,061,524 or increase significantly to £11,212,183 based on referral volume assumptions. If base case rates rise by 9 or 28% estimated total costs of treating skin cancer will increase to £18.1 million and £20.4 million respectively.

**Conclusions:** Skin cancer management costs in Northern Ireland vary from ∼£14.3m to £26.2m depending on diagnostic referral assumptions. Malignant melanoma costs have risen ∼10 fold over the past decade mainly due to chemotherapy costs. Predicted 28% increase in melanoma cases by 2040 would lead to £3.3m of additional referral, diagnostic and treatment expenditures, which with inflation adjustment to a further budget requirement of approximately £6.4 million by 2022 rates.

## Background

Skin cancer is one of the most common forms of cancer in the UK. Concerningly, Northern Ireland has been shown to have had the worst five-year survival for melanoma in the UK^3^. Since the early 1990s, melanoma skin cancer incidence rates have more than doubled (140%) in the UK^4^ and it is expected that in the UK by 2025 and 2040 there will be an increase in malignant melanoma (MM) diagnoses of 9% and 28% respectively^5^. Northern Ireland Cancer Registry figures suggest increases may be greater, with estimates showing expected rates of MM to increase by 28% (by 2025) and 65% (by 2040) in men and 23% (by 2025) and 48% (by 2040) in women^6^. Coupled with this a continuous long-term increase in non-melanoma skin cancer (MNSC) incidence shows no tendency for levelling off ^7^.

The increasing incidence of and mortality from skin cancer has been projected to pose a large financial burden^8^, with estimated to cost the NHS over £180 million in 2020^2^, and the predicted costs of diagnosing and treating cases of Basal cell and Squamous cell cancers were further predicted to increase to ∼£338 to £465 million for 2025^9^. There is therefore a prescient need to understand the costs associated with skin cancer and the triage burden that is associated with the delivery of services for Northern Ireland. Such evidence will prepare policymakers for the economic impact of anticipated changes in the demand for skin cancer services and reignite the significance of efforts to enhance efforts to prevent skin cancer. Policymakers must have evidence of the likely benefits and opportunity costs to inform the review of existing policies and prepare for necessary changes to demand for services which are imminent.

This study examines the cost of illness economic impact of diagnostic and treatment services for skin cancer management in Northern Ireland. We estimate the current and future costs to skin cancer services by 2025 and 2040 of expected disease increases, to provide a basis for policy and planning relative to prevention and control initiatives^10^; the indicative expenditures will inform cost-effectiveness analyses, ground practice optimisation and sustain effective clinical outcomes for patients. This project was initiated within the research objectives of the implementation group for the Northern Ireland Skin Cancer Prevention Strategy^11^.

## Methods

In this paper, we conduct a retrospective cost of illness study to quantify the Northern Ireland Skin Cancer burden, using a health care system perspective. In the analysis, two types of skin cancer were examined. The classifications used were derived from the International Classification of Diseases: malignant melanoma (ICD10: C43), and non-melanoma skin cancer (ICD10: C44). Healthcare costs were considered for all activity that flows from the point of suspicion of skin cancer. In-year costs were reported based on 2018 NHS reference unit costs in UK pounds sterling (UK£)^12^. Inflation indices (RPI) were applied to unit costs obtained where 2018 was not the cost base year. Analyses were conducted in <u>R Version 4.1.2</u>.

We adopted a bottom-up method^1,2^ to specify parameters in a data-driven cost model of skin cancer diagnosis and treatment following the patient pathway, as shown in Figure 1. Patient care attributes were characterised by NICE management guidelines^13^ and supplementary details were validated as by subject experts, including dermatologists, surgeons, oncologists, cancer pathway trackers, cancer service managers, radiology managers, information specialists, regional (NICAN) leads and Cancer Registry staff, who reviewed the structure of the model assumptions to ensure they reflected current practice.

**Figure 1.**
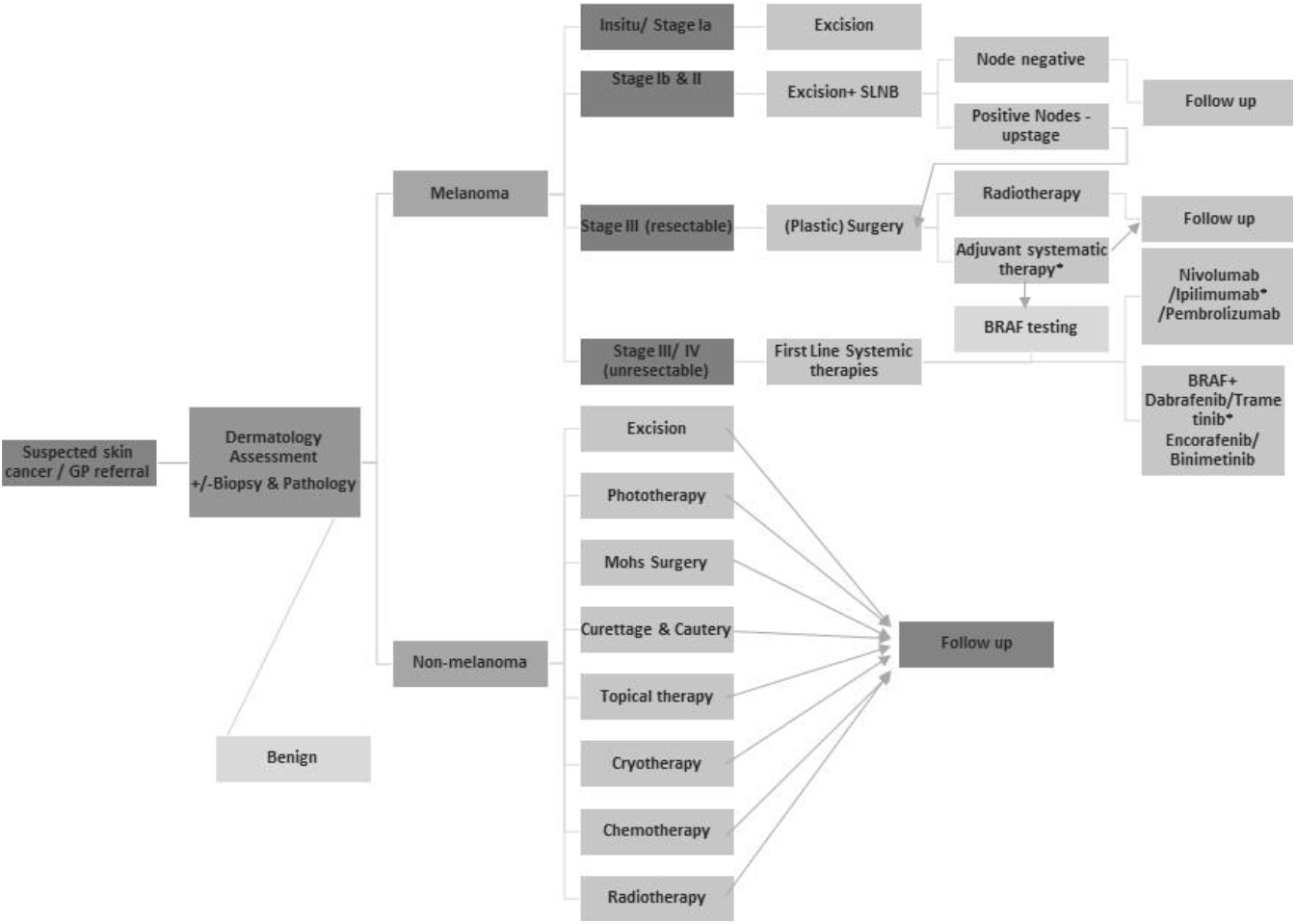
Skin Cancer patient pathways.

In our estimation, we applied Northern Ireland Cancer Registry (<u>NICR</u>) incidence rates of skin cancers (2018), and diagnostic volumes provided by regional health service monitoring teams to calculate the total clinical diagnostic burden including benign cases assessed which flow into subsequent model phases, detailed in Table 1. Data on the proportion of melanoma and non-melanoma cases receiving specified treatments were retrieved from the performance indicators for the registries of the UK and Ireland Association of Cancer Registries (UKIACR)^14^ for 2018, as indicated in the model assumptions in Table 1.

**Table 1.** Model Assumptions.

| Model Attribute | Data | Guidance Notes for Base Case |
| --- | --- | --- |
| Probability of Stage 1 melanoma | 0.715 | Assume of these 63% Stage Ia |
| Probability of Stage 2 melanoma | 0.192 |  |
| Probability of Stage 3 melanoma | 0.064 |  |
| Probability of Stage 4 melanoma | 0.029 |  |
| Cancer conversion rate<sup>29</sup> | 6.8 | UK national data |
| Benign case rate <sup>2</sup><br>(Sensitivity analysis) | 12 per<br>MMSC |  |
| GP referrals | 1 | Per suspected case (19,283 referrals - data supplied by SPPG personal correspondences) |
| Cost per GP visit | £39.23 | NHS Reference Cost 2018/19 |
| Cost per clinical dermatology first appointment (outpatient) | £154 | Assume x1 for all benign, NMSC, and MMSC cases / NHS Reference Cost 2018/19 |
| Multidisciplinary meeting presentation | £123 | Assume 1 for all MMSCs and a further 2 for all SLNB cases / NHS Reference Cost 2018/19 |
| Plastic surgery assessment | £143 | NHS Reference Cost 2018/19 |
| Anaesthetics pre-assessment for SLNB | £106 | Cost per hour – assume a 15-minute appointment |
| Sentinel lymph node biopsy | £3542.65 | Based on commissioned case volume, includes basic surgery excision combined with pathology. Applied in line with NICE |
| Ultrasound Scan | £102 | RD43Z (20 minutes+, with Contrast) – used in follow up Stage Ib/ IIa |
| CT scan | £110 | 3 areas with contrast. Assume x1 at diagnosis and x2 follow-up for all Stage IIc and higher patients. NHS Reference Cost 2018/19 |
| CT scan reporting | £28 | NHS Reference Cost 2018/19 |
| Gene testing<sup>16</sup> | £95 | Based on BRAF testing costs. Applied to all Stage IIc and higher patients. |
| Vitamin D test | £17 | All MMSC patients (NICE) |
| LDH testing | £1 | All Stage 4 MMSC patients |
| **Non-Melanoma Skin Cancer Costs** |  |  |
| Surgical excision | £151 | NHS Reference Cost 2018/19 (JC43C) - Outpatient Minor Skin procedure rate – applied to 0.436 if NMSC cases are based on UKIACR data. **Sensitivity Rate - 0.860<sup>2</sup>** |
| Curettage & Cautery<sup>2</sup> | £180.23 | Applied to 0.075 NMSC cases |
| Phototherapy (per session) | £94 | NHS Reference Cost 2018/19 (JC47Z) – Phototherapy<br>Assume 3 sessions /week x8 weeks, then 1 session every 2 weeks up to 1 year (46 sessions total) – applied to 0.008 of NMSC cases |
| Mohs Surgery<sup>2</sup> | £128 | Applied to 0.004 of NMSC cases |
| Topical therapy | £121.50 | Imiquimod - Apply daily 5 nights /week x6 weeks, apply to the lesion and 1 cm beyond, assess response after 12 weeks<br>- £48.60 for 12 sachet packs or £4.05 per pack - 30 packs required – applied to 0.005 of NMSC cases |
| Cryotherapy<sup>2</sup> | £279 | Applied to 0.031 NMSC cases |
| Radiotherapy | £1002 | NHS Reference Cost 2018/19 SC46Z (Preparation for Simple Radiotherapy with Imaging & Dosimetry, w/ Technical Support x1) @£522 + SC21Z (Deliver a Fraction of Treatment on a Superficial or Orthovoltage Machine x5) @£96. Based on most common reports 35 Gy/#5 fraction regimens<sup>30</sup>, applied to 0.008 NMSC cases as per UKIACR data |
| Chemotherapy | £197.40 | Efudix - usually used once or twice a day for 6 weeks (BAD). Assume max 6 tubes @ £32.90 (NICE). Applied to 0.005 NMSC cases. No procurement cost applied. |
| **Melanoma Skin Cancer Costs** |  |  |
| Surgical excision | £618 | NHS Reference Cost 2018/19 (JC43C) Day case rate for Minor Skin procedures – applied to all Stage 1 MMSCs, with adjustment for SLNB cases |
| Inpatient stay<sup>17</sup> | £3720 | ~33.4% of all referrals will require an inpatient stay of mean 3 days based on Hospital Episode Statistics 2018/19 |
| Radiotherapy | £712 | NHS Reference Cost 2018/19SC50Z (Preparation for Superficial XRT w/Imaging & Dosimetry, w/ Technical Support x1) @£577 + SC21Z (Deliver a Fraction of Complex Treatment on a Megavoltage Machine x1) @£135 - Based on Single-dose radiotherapy<sup>31</sup> of a least 8 Gy for bone metastases. Applied to 0.009 MMSC cases. |
| New Medical Oncology assessment | £208 | NHS Reference Cost 2018/19 - Assume x1 for all chemotherapy and radiotherapy MMSC cases |
| Review Medical Oncology assessment | £196 | NHS Reference Cost 2018/19 - Assume x 1 for each course of chemotherapy + x2 review appointments post-treatment in year 1 |
| Dermatology Review | £136 | NHS Reference Cost 2018/19 |
| Specialist Plastic surgery review | £124 | NHS Reference Cost 2018/19 |
| Clinical Nurse Specialist Health needs assessment | £113/hr | NHS Reference Cost 2018/19 - Assume x2 of 1 hr, 1 at diagnosis and 1 post-treatment. Additional x2 support calls (of 15mins) for all Stage IIc and higher patients. |
| Pembrolizumab<sup>32</sup> | £5260/<br>cycle | 200 mg (IV over 30 minutes, 3 weekly 3 max 1-year adjuvant) - Applied to 0.054 MMSC cases |
| Nivolumab<sup>33</sup> | £5266/<br>cycle | 480 mg (IV over 60 minutes, 4-weekly for max 1-year adjuvant - Applied to 0.028 MMSC cases |
| Ipilimumab<sup>34</sup> | £15,000/<br>cycle | 3 mg/kg (IV over 90 minutes, 3 weekly x 4 cycles only) 3 mg/kg @ £15000 for 200mg/ 40ml vial - assume x1 vial only used approximation of 66.6kg used in lieu of average Europe weight of 155lbs (or 70kg) - Applied to 0.009 MMSC cases |
| Ipilimumab/Nivolumab combination | £15,878/<br>cycle | 3-weekly x 4 cycles then 4 weekly, for a total of 2 years maximum. (3 mg/kg Ipilimumab as above + 1mg/kg Nivolumab - here apply 40mg/4ml vial x2 @439 per vial) - Applied to 0.037 MMSC cases |
| Dabrafenib<sup>35</sup>/Trametinib<sup>36</sup> combination | £5040/<br>cycle | 4-weekly, continuous until disease progression or max 1 year if adjuvant (Dabrafenib 150mg BD as 75mg capsules x 2/dose, 28 caps per pack, £1400 for 7-day supply + Trametinib 2mg once daily, 2mg tablets, £1120 for 7-day supply) - Applied to 0.050 MMSC cases |
| Encorafenib<sup>36</sup>/Binimetinib<sup>37</sup> combination | £7840/<br>cycle | 4-weekly, continuous until disease progression (Encorafenib 450mg OD, 75mg capsules, supplied as 42 capsules, £1400 for 7-day supply + Binimetinib 45mg BD, 15mg tablets, x3/dose, 84 tablets, £2240 for 28-day supply) - Applied to 0.016 MMSC cases |
| Chemotherapy procurement | £2093 | Assume all chemo drugs - Outpatient - NHS Reference Cost 2018/19 - SB10Z Procure Chemotherapy Drugs for Regimens in Band 10 (correspondence Coding Manager BHSCT) |
| New Chemotherapy administration A | £428 | NHS Reference Cost 2018/19 SB12Z - Applied to Pembrolizumab/ Nivolumab regimens |
| New Chemotherapy administration B | £530 | NHS Reference Cost 2018/19 SB13Z - Applied to Ipilimumab |
| New Chemotherapy administration C | £542 | NHS Reference Cost 2018/19 SB14Z - Applied to Ipilimumab/Nivolumab |
| Oral Chemotherapy administration | £328 | NHS Reference Cost 2018/19 SB11Z - Applied to Dabrafenib/Trametinib & Encorafenib/Binimetinib |
| Subsequent cycles chemotherapy | £422 | NHS Reference Cost 2018/19 SB15Z |
| Chemotherapy Helpline contact<sup>38</sup> | £110 | Per hour cost – Band 6 Nurse. Assume 1.28 contacts per patient, 0.5 hours per call for triage, +/- admissions. |
| Vitamin D supplement | £20.70 | Inflation-adjusted annual cost<sup>36</sup>, applied to 12% MMSCs likely to be Vitamin D deficient<sup>39</sup> |
| End of life Unscheduled care in the last year of life Non-Melanoma Skin Cancers | £81,931 | Based on published data 2022<sup>18</sup>, inflation applied |
| End of life Unscheduled care in the last year of life MMSCs | £240,141 | Based on published data 2022<sup>18</sup>, inflation applied |

NOTE: All costs adjusted to 2018 (where this was not the original year of reporting).

### Diagnostic Cost

Following a primary care GP referral, all patients were assessed in secondary care. Diagnostic biopsies are assumed to be a bundled cost within a multi-disciplinary dermatology outpatient new assessment visit cost. For patients with malignant melanoma diagnostic staging costs were included. All MMSCs were assumed to have their case presented once to multi-disciplinary meetings (MDM) and if required a sentinel lymph node biopsy (SLNB) carried out by the recently established service at one hospital trust^15^. For SLNB patients two additional MDM presentations were assumed. The costings for this procedure are solely based on commissioned costs. For all patients with Stage IIc and higher staging, we assumed that a CT scan would be performed at the time of diagnosis. During the baseline assessment of the patient following NICE guidance, the cost of genetic testing, Vitamin D testing, and levels of LDH were included^16^.

### Treatment Cost

To determine the probability of treatment of non-melanoma skin cancer, we used published literature data^2^ regarding excision, phototherapy, topical therapy, and cryotherapy, as well as UKIACR data concerning radiotherapy and chemotherapy use^14^. The study assumed that all the treated cases would receive one review, and 15% of those cases would be reviewed again to re-evaluate the outcome of the treatment.

Case numbers for the service level chemotherapy utilisation data for melanoma patients were unavailable from UKIACR for 2018 for Northern Ireland and were supplemented by data provided by treating clinicians. First-line and subsequent lines of treatment as well as palliative use of chemotherapy were included. Based on NHS reference^12^ and British National Formulary costs, medical oncology assessment and review costs, chemotherapy procurement, delivery, drug reimbursements, and follow-up costs were included. For chemotherapy pre-assessment testing or supportive prescribing, no detailed micro-costs were included. Using Hospital Episode Statistics for inpatient care^17^, we assumed that 33.4% of MMSC patients required an inpatient stay, assuming based on average stay details a stay of 3 days in our study.

### Follow up cost

Based on clinical guidelines a dermatology review for Stage I to IIb melanoma patients is included in the follow-up assessment costs. CT scanning twice in the first year of follow-up and 4 consultant reviews (2 oncology and 2 plastics) were included for patients with Stage IIc and higher staging.

Based on clinical expert opinions, 2 specialist nurse triage/support telephone calls (15 minutes/call) were assumed for all patients with Stage IIc or higher staging. In addition, applying data from previous analyses, inflation-adjusted costs for unscheduled care in the last year of life were added^18^. Follow up costs for prevalent melanoma cases in years 2-5 was based on NICE guidelines^16^.

### Future Cost

To estimate future costs, based on the projected increases of 9% and 28% in melanoma cases^5^, we derived proportionate increases to total diagnostic, treatment and follow-up volumes for malignant melanoma. No changes to background cancer conversion or benign case rates, stage distribution or attendant treatment proportions were modelled.

### Sensitivity Analyses

Given that more than 35% of melanomas are known to be diagnosed outside of the red flag/ two-week wait pathway^19^, we sought to assess the cost implications of complexities of clinical referral pathways and triage in sensitivity analyses. Since there is no definitive evidence of whether these diagnoses arise from urgent and routine dermatology referrals and consequently uncertainty arises in fully characterising the true diagnostic assessment burden. Therefore, sensitivity analyses examine the impact of varying attribute assumptions in two alternate scenarios for diagnostic volumes a) applying a multiplier of 12 benign cases for each melanoma case as previously applied^2^, and b) assuming a diagnostic conversion rate of 6.8%^20^ which is applied to total incidence can provide the volume of cases assessed from which a true diagnosis arises. In addition, we examined alternate rates of surgical excisions of non-melanoma skin cancers. Finally, chemotherapy regimens were assumed to have been fully compliant in the base case, and a 75% utilisation rate was applied in a sensitivity analysis.

## Results

As of 2018, NICR recorded 4142 NMSC and 423 MM cases, an average of 17.5 new patients per trust, per week. A total of £17,024,115 is spent on skin cancer care by healthcare providers. Nonmelanoma skin cancer management costs £1,815,936 (average £438 per case) and melanoma skin cancer costs £12,364,220 (average £29,299 per case). Based on the referral volume data provided by regional planning teams, the base case diagnostic assessment cost (GP referral and dermatology assessment) for cases which will ultimately be benign is estimated to total £2,844,106. The diagnosis of non-MSCs costs £637,868 and the diagnosis of MMSCs was estimated to cost £938,942. In the base case, the total overall cost of diagnostics was estimated to be £3,843,382.

As shown in Table 2, in the base case scenario, total treatment costs for non-melanoma skin cancers amounted to £933,646. The total treatment costs for MMSCs were £9,718,843, of which £8,792,208 was devoted to procurement, administration, and chemotherapy drug use. The attributable costs for access to the chemotherapy helpline by MMSC patients were estimated to be £5878. Therapeutic Vitamin D supplements contributed a further £1050 to the cost.

**Table 2.**
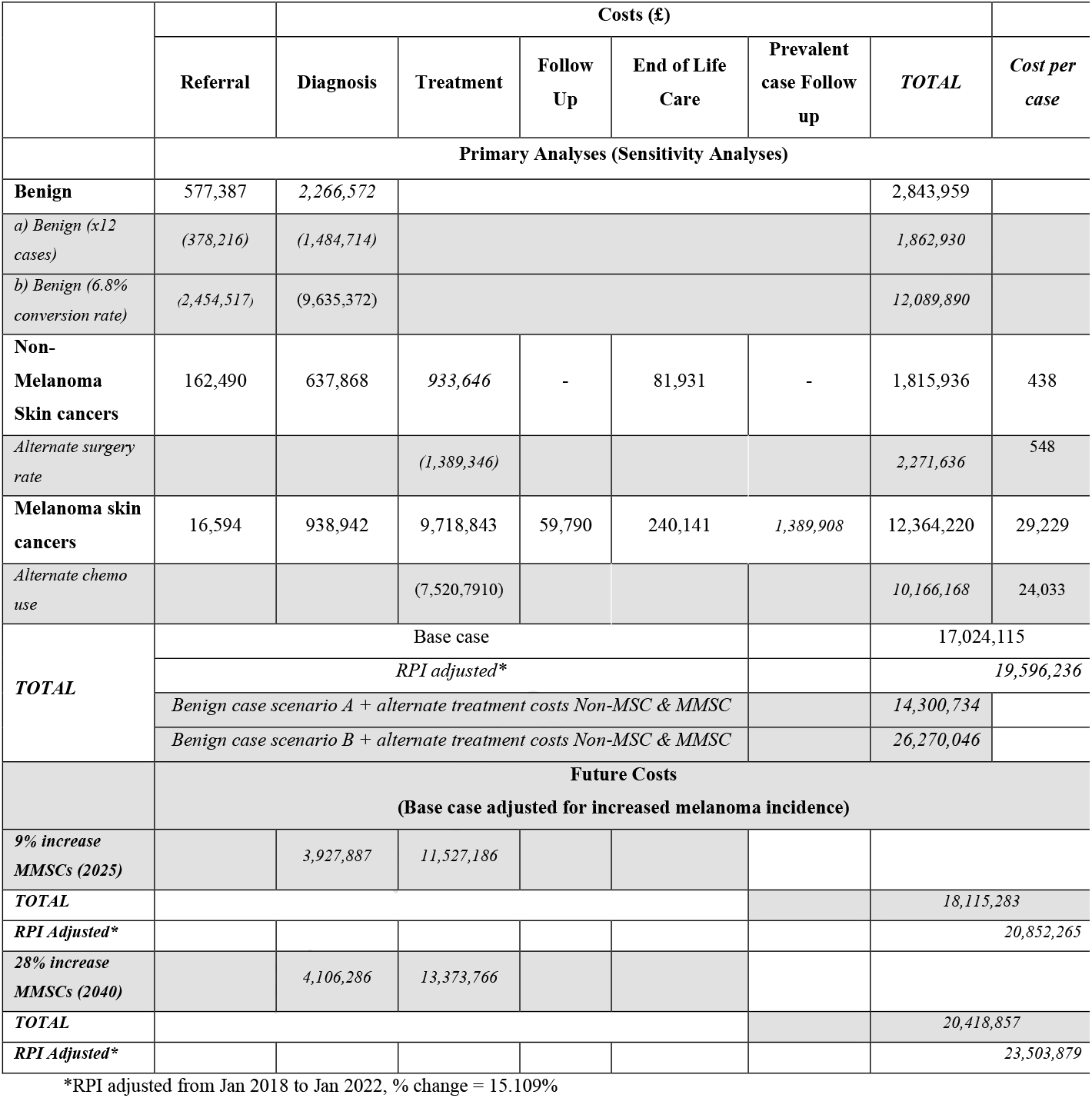
Costs of skin cancer diagnosis, treatment, and care in 2018.

|  |  | Costs (£) |  |  |  |  |  |  |
| --- | --- | --- | --- | --- | --- | --- | --- | --- |
|  | Referral | Diagnosis | Treatment | Follow Up | End of Life Care | Prevalent case Follow up | TOTAL | Cost per case |
|  | Primary Analyses (Sensitivity Analyses) |  |  |  |  |  |  |  |
| Benign | 577,387 | 2,266,572 |  |  |  |  | 2,843,959 |  |
| a) Benign (x12 cases) | (378,216) | (1,484,714) |  |  |  |  | 1,862,930 |  |
| b) Benign (6.8% conversion rate) | (2,454,517) | (9,635,372) |  |  |  |  | 12,089,890 |  |
| Non-Melanoma Skin cancers | 162,490 | 637,868 | 933,646 | - | 81,931 | - | 1,815,936 | 438 |
| Alternate surgery rate |  |  | (1,389,346) |  |  |  | 2,271,636 | 548 |
| Melanoma skin cancers | 16,594 | 938,942 | 9,718,843 | 59,790 | 240,141 | 1,389,908 | 12,364,220 | 29,229 |
| Alternate chemo use |  |  | (7,520,7910) |  |  |  | 10,166,168 | 24,033 |
| TOTAL | Base case |  |  |  |  |  | 17,024,115 |  |
|  | RPI adjusted* |  |  |  |  |  | 19,596,236 |  |
|  | Benign case scenario A + alternate treatment costs Non-MSC & MMSC |  |  |  |  |  | 14,300,734 |  |
|  | Benign case scenario B + alternate treatment costs Non-MSC & MMSC |  |  |  |  |  | 26,270,046 |  |
|  | Future Costs<br>(Base case adjusted for increased melanoma incidence) |  |  |  |  |  |  |  |
| 9% increase MMSCs (2025) |  | 3,927,887 | 11,527,186 |  |  |  |  |  |
| TOTAL |  |  |  |  |  |  | 18,115,283 |  |
| RPI Adjusted* |  |  |  |  |  |  | 20,852,265 |  |
| 28% increase MMSCs (2040) |  | 4,106,286 | 13,373,766 |  |  |  |  |  |
| TOTAL |  |  |  |  |  |  | 20,418,857 |  |
| RPI Adjusted* |  |  |  |  |  |  | 23,503,879 |  |

\*RPI adjusted from Jan 2018 to Jan 2022, % change = 15.109%

During the first year after the diagnosis of skin cancer, the estimated total cost of follow-up for complex melanoma cases, was £59,790. Follow up costs for cases in years 2-5 following diagnosis was estimated to be £1,389,908.

For patients with non-melanoma and melanoma skin cancers, an additional £322,072 would also be incurred for the unscheduled emergency admission care for those in the last year of their lives.

### Sensitivity Analyses

In sensitivity analysis for alternate diagnostic volumes to determine the benign case rates, we examined two alternative parameter conditions, firstly a scenario where, as previously applied, 12 benign cases were treated per MMSC case. Making this assumption leads to an estimated referral volume of ∼9641 patients, in so doing the ensuing GP referral costs reduced to £378,216. However, by applying a national diagnostic conversion rate of 6.8% to estimate the true diagnostic volume, this would result in 62,567 patient triage assessments annually (240 per week in each trust), which would result in an annual GP referral cost of £2,454,517 for this scenario. The consequent diagnostic assessment of cases applying these rates for alternate diagnostic scenarios for true dermatology assessment volume would result in diagnostic costs for ultimately benign cases ranging from £1,484,714 and £9,635,372. Therefore, our sensitivity analyses, show that the overall diagnostic cost could either reduce to £3,061,524 or increase significantly to £11,212,183, with the former scenario being shown to be implausible based on the existing referral volume data.

Two costing scenarios for the treatment of NMSCs were examined, using different rates for surgical excision. The sensitivity analysis applied a higher proportion of patients receiving surgical excision for non-melanoma skin cancers, which led to total costs for non-melanoma skin cancer treatment were £1,389,346.

By varying the level of compliance with chemotherapy for melanoma skin cancers, we lowered the cost for procurement, administration and use of chemotherapy drugs by 25%, assuming that overall use was 75% of that of the base case. By doing so treatment costs for MMSCs were lowered by £2,198,052 to £7,420,791.

Assuming both these changes to treatment scenarios were combined in effect simultaneously, the joint impact on costs would see treatment costs lower to £8,910,137 from £10,652,490.

### Predicted Increases in Melanoma Rates

The total costs of treating skin cancer increase to £18,115,283 if the base case melanoma rates increase by 9%. However, if the base case melanoma rates increase by 28%, the total costs will rise to £20,418,857.

Nevertheless, the base case costs will increase to £19,596,236 if RPI adjustments are applied to generate a 2022 cost correction (a 15.1% increase). Details of the potential future cost impacts of an increase in melanoma rates are provided in Table 2.

## Discussion

This study provides the first estimates of the cost of illness in skin cancer management for Northern Ireland which range from ∼£17m to £26.2m in the base case and sensitivity analyses.

The variation in estimates is due to the assumptions made regarding referral and diagnostic exclusion of cases expected to be benign. The sensitivity analyses rely on conversion data primarily sourced from two-week wait data, but this pathway does not diagnose most melanomas^19^. The actual assessment volume is obscured by dermatology referral overlaps and data limitations, hindering the full characterization of benign cases and potentially consuming significant hospital resources. Given the ∼£10m cost disparity and the awareness that 53% of melanoma diagnoses do not involve Red Flag indicators, further refinement of data pathways is necessary for an accurate assessment of skin cancer diagnostic costs^19^. This highlights the need to improve referral capture and capacity planning in anticipation of rising demand. This information is valuable for decision-makers seeking to enhance data intelligence.

When last evaluated a decade ago the expected costs per case of malignant melanoma was ∼£2600 and the expected costs of nonmelanoma skin cancer was ∼£890-£1200^2^. During that period there has been a rapid transformation in the available range of therapeutics for melanoma care. Our results show that the cost per case for malignant melanoma has now risen to ∼£29k – a more than 10-fold increase in the cost of care over the period. Our estimates for the management of non-malignant care a lower than those in the previous study. The relatively low cost (£933k-£1.3m) of treating non-melanoma skin cancers should be interpreted with caution since non-melanoma incidence data are recorded for only the first case in each individual, and true costs are therefore likely to be higher.

Furthermore, our assumptions on the practice patterns are based on previous assumptions, whilst in real world settings treatment for solitary lesions compared to field directed treatments for multiple lesions is constantly evolving^21^. In addition, we note that the former study assumptions for follow-up volume and inpatient care are likely to cause the divergence in estimates. Whilst our estimates for non-melanoma follow-up are conservative, we did not find evidence within HES data of inpatient care for non-melanoma cancers – there are three other reference classes for ‘skin’ noted – as such no definitive method to delineate or retrieve accurate relevant data for non-melanomas was available.

The implied costs from the simplified assumption of one follow-up visit (based on clinical consensus) broadly accord with other reports^22^ which suggest that the median number of day cases for non-melanoma care was 1.5, however, future work should seek to consolidate true inpatient activity in for the non-melanoma setting.

Chemotherapy use within the melanoma cases in our study necessitates 90% of the overall cost of malignant melanoma management in the base case, assuming that real-world compliance with the regimens is lower, as in the sensitivity analyses this reduces by ∼£2.1m. In addition, clinical coding practice on procurement charges may be revised and this may deliver further cost reductions. Whilst not explored herein the resource burden of adverse event management for immunotherapy use in melanoma can add up to £2835-£2860 per case for the management of immunotherapy induced colitis^23,24^, however, the prevalence of severe side effects is low^25^ and in these instances, planned therapies would be delayed or omitted. As such the additional adverse event costs are unlikely to significantly outweigh omitted drug costs, and the net effect may realize a reduction in the overall cost.

Future costs predictions have been conservatively estimated for the expected rise in melanoma rates assuming only the additional referral and diagnostic costs, yet a 9% increase in melanoma cases, would be expected to add a further ∼£1.09m to management costs, which ultimately would accrue inflationary rises leading to an additional cost, relative to the base case which is more likely to be in the range of £3.8 million by 2025.

By 2040 the predicted additional 28% of melanoma cases, would result in an additional £3.39m of referral, diagnostic and treatment costs, which with inflation added to 2022 rates, would see an additional budget requirement of ∼£6.47 million notwithstanding any changes to practice or stage distribution of disease. Critically therefore, it is important given these expected increases in melanoma incidence to consider the crucial mediating effects of interventions such as banning indoor tanning accompanied by a public health campaign, which has the potential to offset the increase in incidence and provide ∼4.8% reduction in melanoma cases^26^.

### Limitations

We were unable to characterise the clinical follow-up regimens for those with unknown staging (n=118). However, the treatment volume and case data provided by clinical oncology teams included treating all cases. As such we do not believe there are underestimates of re-treated and palliative patient chemotherapy volumes, however, prevalent cases follow-up from year two onwards was not fully costed for those with unknown stage at diagnosis. In addition, we did not characterize the costs for those with brain metastases, this would lead to the results presented reflecting an under-estimate of the overall melanoma skin cancer management costs.

Furthermore, we acknowledge that 5% of cutaneous squamous cell carcinoma cases may present as either locally advanced or metastatic^27^ and based on recent advances may now access cemiplimab for routine use^27^, the cohort presented will not have been afforded this option and these costs are not reflected in the data. Other areas of the UK have also introduced the use of brachytherapy to treat certain types of basal cell or squamous cell skin cancers^28^, however, data on the volume of use in the NI setting are not included here due to the paucity of data on access to these treatments. Future research should seek to include this activity in their reporting.

## Conclusion

The estimated costs for the diagnosis, treatment and care for skin cancer range from ∼£12.9m to £26.2m. The modest cost of £933k - £1.3m for the management of non-melanoma skin cancers is outweighed by the management of the ultimately benign case assessment costs. The cost per case for malignant melanoma has now risen to ∼£29k a more than 10-fold increase in the cost of care over the last decade. The predicted increase in melanoma cases by 2040 of 28% would result in an additional budget requirement of more than ∼£3.3 million, without attendant increases in capacity such demand would undoubtedly lead to delays in diagnosis and care.

## Data Availability

All data produced in the present work are contained in the manuscript

## Acknowledgements

Expert involvement at the initiation of this project was provided by Dr Olivia Dolan, Dr Collette McCourt, Dr Bryan Murphy, Dr Bode Oladipo, Dr Joe Houghton, Dr Andrea Corry, Lisa Ranaghan, Maria Wright, Ciara Toal (Belfast Health and Social Care Trust) and by Sheena Stothers, Roisin Hill, Mary Jo Thompson, Karen Robinson (South Eastern Health and Social Care Trust). During analysis, advice was provided by Prof Ed Wilson (Exeter University). Data support provided by Sinead Hawkins and Helen Mitchell (Northern Ireland Cancer Registry), and resource data for health service costs was also provided by Ciara Shivers (DHPSSNI). Coding clarifications were provided regarding chemotherapy procurement by Rosemary Hutton (Belfast Health and Social Care Trust). Sarah Donaldson – Strategic Performance and Planning Group / Public Health Agency NI, provided support on referral volumes.

